# Script Generation as an Efficient Measure of Cognition and Everyday Function in Older Adults

**DOI:** 10.64898/2025.12.18.25342597

**Authors:** Melissa Rosahl, Marina Kaplan, Moira McKniff, Anna Callahan, Riya Chaturvedi, Tania Giovannetti

## Abstract

**Objectives:** A Script Generation Task (SGT), requiring participants to verbally describe the steps of everyday activities, was investigated as an efficient tool to detect mild functional difficulties and mild cognitive impairment (MCI).

**Methods:** 83 participants (n=57 HC; n=26 MCI) completed a SGT, performance-based test of everyday function, cognitive tests, and questionnaires. SGT responses were transcribed and scored by human raters and automated text analysis.

**Results:** Participants with MCI generated fewer SGT steps in a shorter amount of time and more pronouns relative to nouns, reflecting less specificity. Performance on the SGT was associated with cognitive tests of episodic memory, performance-based tests of everyday function, and questionnaires regarding everyday functioning.

Conclusions

The SGT holds promise as a highly efficient measure of mild cognitive and functional difficulties in older adults.

## INTRODUCTION

As cases of Alzheimer’s disease and Alzheimer’s disease related disorders (AD/ADRD) are predicted to triple by 2050 (Nichols et al., 2022), there is a heightened demand for sensitive, cost-effective diagnostic assessments. Conventional assessment includes comprehensive neurological evaluation, neuroimaging, and invasive biomarker testing of AD/ADRD, which imposes a large financial burden on patients, caregivers, and healthcare systems. With advances in pharmaceutical treatments, inexpensive and streamlined early detection of AD/ADRD, at the stage of mild cognitive impairment (MCI), for example, is crucial. Because some mild cognitive decline is common in advanced age, it is especially important to be able to detect early pathological changes that confer risk for functional decline and disability. Functional impairment is one of the most devastating consequences associated with AD/ADRD, leading to depression, low quality of life, and financial and caregiver burden. The ability to effectively and efficiently detect risk for future functional disability could facilitate efficient workflows for the delivery of pharmaceutical and lifestyle-changing interventions (Sabbagh et al., 2020). The goal of the current study was to investigate the potential for a Script Generation Task (SGT), which requires verbally stating the steps required to perform everyday tasks, as a tool to identify older adults at risk for AD/ADRD and functional impairment.

Functional decline co-occurs with cognitive decline, with mild functional difficulties noted early in the course of neurodegenerative disease. For example, on self-report measures and performance-based measures of everyday function, a greater number of subtle errors and inefficiencies are reported by older adults relative to younger adults and by people with MCI versus older adults with healthy cognition (McKniff et al., 2025). However, there are drawbacks to questionnaires (e.g., potential for bias) and performance-based tests (e.g., time-intensive).

SGTs, especially when scored with automated methods like natural language processing (NLP), may offer a more efficient measure to quickly screen older adults at risk for AD/ADRD and everyday function difficulties.

Speech and language have been studied over several decades for early detection of neurodegenerative disease. Speech assessments have become more popular with automated coding of recorded language samples and focus on the motoric and acoustic aspects of verbal responses (Martínez-Nicolás et al., 2021; Themistocleous et al., 2020). However, speech metrics may be biased by age, gender, emotional state, and even microphone/recording properties.

Analyses of semantic and linguistic features of language samples have an even longer history, with a large literature including human scoring of lexical semantic difficulties on standardized tasks (e.g., anomia, reduced verbal fluency) (Eyigoz et al., 2020). Language changes are identified in the very early stages of decline, often before significant cognitive deficits and functional disability arise (Robin et al., 2021). Several studies have used automated natural language processing (NLP) analyses to automate the quantification of linguistic features of complexity and specificity and shown significant and reliable differences between people with healthy cognition versus AD and healthy cognition versus MCI. These studies have shown that groups with greater cognitive impairment show more pronoun use (i.e., less specific language) and lower measures of syntactic complexity and readability (Mueller et al., 2018; Nyongesa et al., 2025). More recently, artificial intelligence (AI) tools, such as large language models (LLMs) have been used to identify linguistic markers of impairment. Researchers have predicted Mini-Mental Status Exam scores from language samples using generative AI (Agbavor & Liang, 2022b, 2022a; Haulcy & Glass, 2021). Several have produced an end-to-end computational diagnostic model of AD based on spoken language (Agbavor & Liang, 2022a; Zhang et al., 2025).

These results could revolutionize dementia screening and assessment, with potential for efficiently identifying large numbers of older adults for cost-saving prevention therapies. The development of at-home cognitive screening assessments, in-clinic speech analysis applications, and secondary analyses of physician visit notes are just some of the proposed solutions to move forward (Du et al., 2024; Konig et al., 2018; Nyongesa et al., 2025; Robin et al., 2021; Sabbagh et al., 2020). While not yet tested on a large scale, these technological interventions hold great potential for both clinicians and the public.

Much of the current literature using automated speech and language analysis for early detection relies on picture description (Cookie Theft Description Task) or unstructured conversations (Agbavor & Liang, 2022b; Cao et al., 2024; Ding et al., 2024; Eyigoz et al., 2020; Mueller et al., 2018; Nyongesa et al., 2025). Such unstructured tasks lack constraints, which allow for compensatory strategies, potentially reducing their sensitivity to detect very mild difficulties. For example, word finding difficulties could be masked by circumlocutory descriptions. Additionally, picture descriptions and conversation may fail to capture cognitive abilities that are closely linked to everyday function, such as sequencing and the ability to sustain attention to achieve a specific goal. To address these limitations, we turned to script generation as an alternative task to elicit language samples for early detection of AD/ADRD.

Scripts are semantic representations of everyday tasks that include information regarding the sequence in which task steps are performed to achieve a larger goal (Funnell, 2001; Grafman et al., 1991; Roll et al., 2019). For example, the script for the activity “leaving the house” may involve 1) finding one’s house and car keys, 2) putting on a pair of shoes, 3) walking outside, and 4) locking the door. Scripts, and the ability to access and activate script representations are crucial for successful completion of everyday tasks (Abbott et al., 1985). The Goal Control Model of everyday function, which was developed to explain the breakdown of everyday activities in dementia, posited that the degradation and/or premature decay of scripts (i.e., goals) leads to failures in completing task steps in everyday life (e.g., omission errors) (Giovannetti et al., 2021). Knowledge degradation and premature decay may be measured by conventional cognitive tests of declarative memory, including tests of semantic and episodic memory. By contrast, executive functions are critical for the efficient and smooth enactment of scripts (i.e., control); when executive functioning is impaired, everyday task steps are performed inefficiently. Steps are mis-sequenced, repeated, or performed imprecisely (e.g., commission errors).

Studies of SGTs have shown that when compared to healthy controls, participants with dementia generate fewer task-relevant steps, and produce more off-task events and sequence errors (Grafman et al., 1991; Roll et al., 2019). In a sample of participants with Parkinson’s dementia or AD (Roll et al., 2019), Roll and colleagues (2019) showed that SGT scores were associated with the accomplishment of task steps on a performance-based test of everyday action, such that participants who generated more essential and task-related steps on the SGT also made fewer task omission errors on the test of everyday action. Furthermore, SGT scores of essential and task-related steps were significantly associated with cognitive tests of episodic memory but not tests of executive function (Roll et al., 2019).

The current study investigated the potential utility of a SGT as a measure of cognition and everyday function to identify older adults at risk for dementia. Both human-coded and automated analyses of language features were performed. We reasoned that the results of human coding could be used to inform predictions for future studies using NLP and other automated language analyses.

The present study had three aims. First, to investigate whether performance on SGTs would significantly differ between people with healthy cognition versus mild cognitive impairment (MCI), who are known to differ in subtle cognitive and functional abilities and risk for dementia. We hypothesized that participants with MCI would generate fewer steps than those in the healthy cognition group. These predicted outcomes are supported by previous research on AD patients and those with healthy cognition (Grafman et al., 1991; Roll et al., 2019). Inclusion of the MCI group extends the extant literature. The second aim was to investigate the relations between performance on the SGT and cognitive tests of episodic memory and executive function. According to the Goal Control model, generation of task steps should be associated with higher episodic memory scores. Measures of script efficiency, such as the generation of unrelated, commentary words and total errors, were predicted to be associated with measures of executive function. Similar results were found in a study of participants with dementia (Roll et al., 2019).

The third aim was to examine relations between the SGT and actual performance of everyday tasks as measured with the Naturalistic Action Task (NAT), as well as self-reported functioning in everyday life. Based on prior work showing script representations are essential for task performance (Abbott et al., 1985; Funnell, 2001; Grafman et al., 1991), we predicted that there would be a significant positive association between verbal script generation and NAT performance. We also predicted that higher performance on the SGT would be associated with higher independence, and lower scores, on all functional activity questionnaires. If supported, the third aim would show the promise of Script Generation as an efficient and objective measure of mild functional difficulties in older adults.

## METHOD

### Recruitment

Participants were recruited as part of a parent study focused on understanding everyday function in older adults (R01AG062503). Recruitment took place in the Philadelphia community (senior centers, the Temple Center for Lifelong learning, local senior apartments, and churches) through flyers and outreach events and through referrals from local neurology departments.

### Participants

Participant inclusion criteria for the parent study were 1) community-dwelling; 2) native English speaker; 3) age 65 years of age or older; 4) availability of a study partner/informant knowledgeable of the participant’s everyday functioning; 5) no history of neurological disease or disorder other than mild dementia; 6) no current psychiatric disorder; 7) no current metabolic or systemic disorder. Eligibility was determined by phone screening and confirmed by measures during the testing session. For the current study, participants who self-reported speaking English as a second language (ESL) on the demographic questionnaire were excluded from the primary analyses.

### Procedure

Participants came to the lab for a 3–4-hour visit. During the visit, participants completed informed consent, the Naturalistic Action Task, followed by cognitive testing, the Everyday Action Script Generation Task, and questionnaires. Participants received a $50 gift card for their time. All study procedures were conducted in compliance with the Temple University Institutional Review Board (IRB) policies on the ethical conduct of human research (IRB protocol #23116).

### Cognitive Testing

Cognitive testing was used to classify participants’ cognitive abilities (e.g., Healthy Cognition vs. MCI; Aim 1) and in correlation analyses with SGT scores (Aim 2). Participants were administered two tests from four different cognitive domains to evaluate Jak/Bondi actuarial criteria (Jak et al., 2009) and clinical criteria originally proposed by Peterson to classify MCI (Petersen, 2004). Specific tests were selected from the Calibrated Neuropsychological Normative System (CNNS), which allows for demographic adjustments (age, sex, education level, estimated IQ) (Schretlen et al., 2009). For correlation analyses, demographically adjusted T-scores were averaged to compute composite scores for episodic memory (Hopkins Verbal Learning Test-delayed free recall; Brief Visuospatial Memory Test-Revised – delayed free recall) and executive function (Trail Making Test-Part B; Digit Span Backwards).

### Naturalistic Action Task

The Naturalistic Action Test (NAT), also referred to as the “Real Kitchen Task,” is a performance-based test of everyday action with strong psychometric properties (Giovannetti et al., 2007, 2019; Rycroft et al., 2018; Schwartz et al., 2002; Seligman et al., 2013). For the present study, the NAT was used in correlation analyses with the SGT scores to evaluate Aim 3. The NAT requires participants to complete a breakfast and lunch task using real objects that are set on the tabletop in a standardized arrangement. Standardized instructions directed participants to work as quickly and efficiently as possible without making errors. Before each trial, participants were asked to repeat the instructions to ensure comprehension. These procedures were used in prior publications for the NAT. Similar to the procedures of the SGT, participants were not given feedback during the NAT and were simply told to “Do what you think is best” if they asked questions while completing the tasks.

Performance was video recorded for review by coders blind to participant details. Three performance measures were obtained (Giovannetti et al., 2007, 2019; Rycroft et al., 2018; Schwartz et al., 2002; Seligman et al., 2013): 1) completion time - number of seconds was recorded from the time of the first movement to when the participant indicated that they were finished; 2) accomplishment steps - were recorded as the number of completed steps during the task (totals: Breakfast-13; Lunch-20); and 3) errors - such as reaching or touching an object without using it, completing extra steps, and motor quality errors were recorded.

### Participant Questionnaires of Everyday Function and Change

Participants completed surveys regarding everyday functioning, including the Everyday Cognition Scale (ECog), Functional Activities Questionnaire (FAQ), and the Instrumental Activities of Daily Living-Compensation (IADL-C) questionnaire. A higher total score for each questionnaire indicated more everyday functioning difficulties and change over time. The ECog assessed decline over the past decade across twelve everyday abilities, such as remembering where you have placed objects, with total scores ranging from 12 to 48. The FAQ included ten activities related to cognitive problems, with zero reflecting the most independent and 30 reflecting the most dependent individual. The IADL-C evaluated independence in completing 27 daily activities on a scale from one to eight, with totals ranging from 27 to 216. Total scores for each questionnaire were used in correlation analyses to evaluate Aim 3.

### Everyday Action Script Generation Test (SGT)

The Everyday Action Script Generation Test required participants to describe the steps that they would take to complete a series of four everyday tasks (see Table 1). The examiner followed a script to deliver instructions (See Supplement A). The SGT started with the examiner providing an example (tooth brushing) to ensure that the task directions were clear and to calibrate the participant to the level of detail requested. After the example, the participant was asked if they had any clarification questions before the first item was administered. Once the participant began an item, they were not provided with any specific instructions and were told to simply “Say whatever you think is best.” If the participant exceeded three minutes in response to any of the items, the examiner would stop them and record the end time. The SGT is a previously validated measure in distinguishing healthy cognition from AD (Roll et al., 2019).

**Table 1:** Everyday Action script Generation Task Instructions.

| Item | Verbatim Instructions |
| --- | --- |
| Sample | If I ask you to tell me all of the steps you would to brush your teeth, you could say:<br>Open toothpaste. Put paste on brush. Turn on water. Wet toothbrush. Put toothbrush in mouth. Brush teeth. Rinse mouth. Rinse teeth. Turn off water. |
| Item 1 (Toast) | Tell me all of the steps that you would do to make toast with butter and jelly. |
| Item 2 (Coffee) | Tell me all of the steps that you would do to prepare instant coffee with cream and sugar. |
| Item 3 (PBJ) | Tell me all of the steps that you would do to make a sandwich with peanut butter and jelly. |
| Item 4 (lunchbox) | Tell me all of the steps that you would do to prepare a lunchbox with a sandwich, a snack, and a drink. |

NOTE: Each item in the first column corresponds to one of the four SGT items. Verbatim Instructions were verbally stated by the examiner at the beginning of each item.
ABBREVIATIONS: PBJ = peanut butter and jelly sandwich

All responses were recorded using a tape recorder, and audio files were uploaded to a secure computer for storage. Audio files were manually transcribed and timestamped by trained undergraduate research assistants. Transcriptions were coded by research assistants blind to participant’s cognitive status following procedures outlined below. Coding questions were brought to a consensus meeting for review and discussion.

### Human Coding of Script Generation Task

Coders reviewed SGT audio files and transcripts and recorded the following scores. An overview of the human coding process instructions is provided in Supplement B.

> Total Time to Completion – Using the audio file, the time the participant spent generating their response after each prompt was recorded in seconds.

> Total Essential Steps – The coders indicated the number of steps from a predetermined list that the participant reported. (See Supplement C)

> Total Non-essential Steps – The total number of additional steps reported that were judged to be related to the task but not included in the list of essential steps. A non-essential step was a step related to the task, but not necessary for completion. These could be preparatory or cleanup steps. An example of a non-essential step is when a participant says they would clean the dishes that were used while making a sandwich.

> Off-task Steps – The total number of steps unrelated to the task. If the participant described how to make toast with butter and jelly, an off-task step would be to phone their daughter to ask how she is doing.

> Total Words – The total number of words generated after each of the prompts.

> Total Script Errors – The total number of lexical substitution, sequence, and perseveration errors. A lexical substitution error was defined as substituting one word for another. For example, the participant may say the word “milk” instead of “cream.” Perseveration errors were defined as the repetition of steps, sub-steps, or words. An example of perseveration could be the following, “I would add sugar, then cream, then add sugar.” Finally, sequence errors were defined as steps in the wrong order. Coders used a predefined sequence of steps to determine sequence errors. For example, adding butter or jelly to bread before toasting was considered a sequence error, but there were no sequential constraints regarding the order of adding butter and jelly.

> Total Commentary Words – The number of words used in discussing ancillary information after each prompt. Commentary was defined as narration that is not relevant to the prompt. Unlike off-task steps, commentary are observations or reactions, rather than actions to take (e.g., “That reminds me, I need to go to the grocery store.). Errors within commentary text were not counted.

### Automated Coding of Script Generation Task

The SGT transcripts were stripped down to participants’ responses and saved as text files labeled by group. Examiner’s instructions, timestamps, and laughter or actions in parentheses were removed. Using the R package, spacyr, we parsed and cleaned the corpus, tagging parts of speech (POS). Punctuation, symbols, and spaces were also removed.

### NLP Measures

Several packages in R (e.g., Quanteda, quanteda.textstats, spacyr, tidyverse) were used to obtain the following variables:

> Pronoun-Noun Ratio-The number of pronouns divided by number of nouns

> Lexical Diversity (Type-Token Ratio (Templin, 1957))-number of token types (unique words) divided by number of tokens (words)

> Lexical Density (Johansson, 2008)-Proportion of lexical items (nouns, adjectives, adverbs, verbs), also called “open-class words”, to total words

> ο Spacy POS tags: NOUN, ADJ, ADV, VERB

> Lexical Sophistication (LAUFER & NATION, 1995)- percentage of “uncommon” words; defined using the NGSL v1.2 of 2,809 most common English words.

### Statistical Analyses

All score distributions were tested for normality before analyses. To evaluate Aim 1, a one-way between-group ANOVA was performed comparing MCI vs Healthy Cognition groups on the Script Generation Test scores. To evaluate Aim 2, correlation analyses were used to examine relations between Script Generation Test scores and conventional cognitive tests of episodic and executive functioning. Finally, Aim 3 was evaluated using correlation analyses examining relations between Script Generation Test scores, NAT scores, and self-reported questionnaire measures. All analyses were performed using R 4.4.3; R studio 2024.12.1+563.

## RESULTS

### Participant demographics and score distributions

Participants included 91 older adults of which 8 participants reported English as a Second Language (ESL) and were not included in the final analytic sample of 83 participants. Of the 83 participants, 57 were classified as having Healthy Cognition (HC) and 26 with MCI. As shown in Table 2, HC and MCI groups did not significantly differ in any demographic category.

**Table 2:** Demographic data of the study sample.

|  | Healthy Cognition<br>(n=57) | MCI (n=26) | Kruskal-Wallis<br>Chi Square<br>(df=1) | ESL Group (n=8) |
| --- | --- | --- | --- | --- |
| Age (years) | 72.9 ± 5.4 | 75.2 ± 8.2 | 0.72*, p=0.39 | 72.9 ± 6.6 |
| Sex (% Female,<br>n) | 70% (40) | 62% (16) | 0.6, p=0.44 | 63% (5) |
| Race (% , n) | 61% (35) White | 42% (11) White | 3.53, p=0.06 | 50% (4) White |
|  | 37% (21)<br>Black/African<br>American | 46% (12)<br>Black/African<br>American |  | 25% (2) Asian |
|  | 1.8% (1) Asian | 7.7% (2) Asian |  | 13% (1) Native<br>Hawaiian/Pacific<br>Islander |
|  |  | 3.8% (1) American<br>Indian/Alaskan<br>Native |  | 13% (1)<br>Unknown/Not<br>Reported |
| Calibrated IQ | 115.51 ± 11.83 | 114.42 ± 12.83 | 0.29, p=0.59 | 101 ± 9 |
| Education (years) | 16.23 ± 2.65 | 15.38 ± 3.26 | 1.31, p=0.25 | 18 ± 1.85 |
| Episodic Memory<br>Composite T-score | 49.18 ± 6.64 | 33.15 ± 6.57 | 44.74, p <0.0001 | 52.81 ± 8.08 |
| Executive<br>Function<br>Composite T-Score | 51.85 ± 7.5 | 46.73 ± 11.12 | 3.375, p=0.07 | 51.56 ± 12.07 |

NOTE: All participants reported non-Hispanic/non-Latino ethnicity. Episodic memory and executive function composite scores are shown as described in section 2.3.1.

Normality was determined by evaluating each variable’s distribution on a histogram.

Essential steps were positively skewed; non-essential steps, commentary words, and total errors were negatively skewed. Non-parametric, Kruskal-Wallis ANOVAs and Spearman rank-order correlations were performed, because the data were not normally distributed.

### Aim 1: HC vs. MCI differences on the SGT

Results from the human coding of SGT responses are shown in Table 3. Consistent with our hypotheses, participants with MCI generated significantly fewer essential steps and non-essential task steps on the SGT. Those in the HC group also took significantly more time to complete the SGT task. The groups did not differ on the total words generated, total errors, or total commentary words generated on the SGT. Groups did not differ on total errors, even when analyzed as a dichotomous variable (e.g., 0 errors vs. 1 or more errors).

**Table 3:**
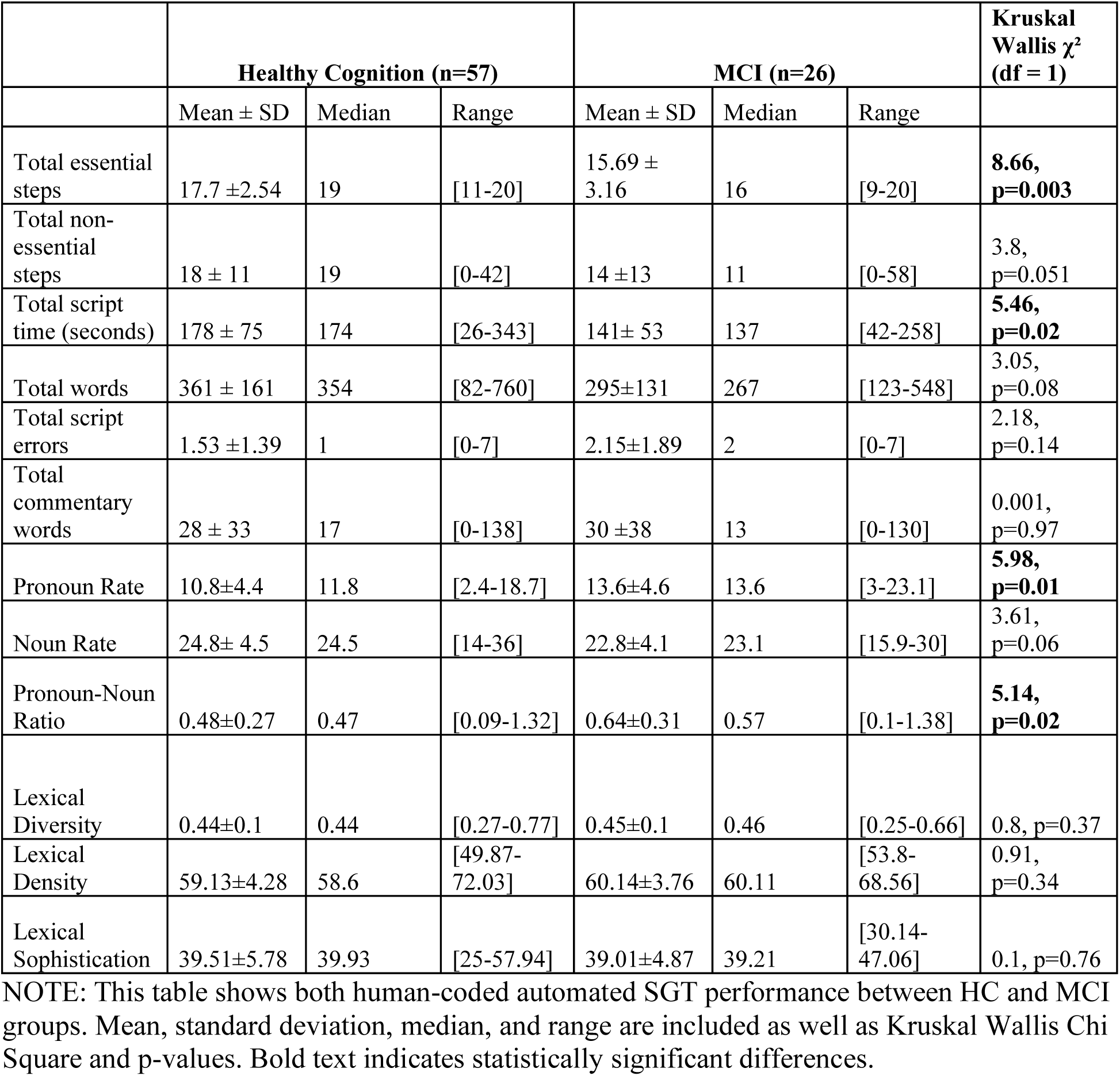
Script generation performance measures (m, SD, median, range)

| | Healthy Cognition (n=57) | | | MCI (n=26) | | | Kruskal Wallis $\chi^2$ (df = 1) |
| --- | --- | --- | --- | --- | --- | --- | --- |
| | Mean ± SD | Median | Range | Mean ± SD | Median | Range | |
| Total essential steps | 17.7 ± 2.54 | 19 | [11-20] | 15.69 ± 3.16 | 16 | [9-20] | **8.66, p=0.003** |
| Total non-essential steps | 18 ± 11 | 19 | [0-42] | 14 ± 13 | 11 | [0-58] | 3.8, p=0.051 |
| Total script time (seconds) | 178 ± 75 | 174 | [26-343] | 141 ± 53 | 137 | [42-258] | **5.46, p=0.02** |
| Total words | 361 ± 161 | 354 | [82-760] | 295 ± 131 | 267 | [123-548] | 3.05, p=0.08 |
| Total script errors | 1.53 ± 1.39 | 1 | [0-7] | 2.15 ± 1.89 | 2 | [0-7] | 2.18, p=0.14 |
| Total commentary words | 28 ± 33 | 17 | [0-138] | 30 ± 38 | 13 | [0-130] | 0.001, p=0.97 |
| Pronoun Rate | 10.8 ± 4.4 | 11.8 | [2.4-18.7] | 13.6 ± 4.6 | 13.6 | [3-23.1] | **5.98, p=0.01** |
| Noun Rate | 24.8 ± 4.5 | 24.5 | [14-36] | 22.8 ± 4.1 | 23.1 | [15.9-30] | 3.61, p=0.06 |
| Pronoun-Noun Ratio | 0.48 ± 0.27 | 0.47 | [0.09-1.32] | 0.64 ± 0.31 | 0.57 | [0.1-1.38] | **5.14, p=0.02** |
| Lexical Diversity | 0.44 ± 0.1 | 0.44 | [0.27-0.77] | 0.45 ± 0.1 | 0.46 | [0.25-0.66] | 0.8, p=0.37 |
| Lexical Density | 59.13 ± 4.28 | 58.6 | [49.87-72.03] | 60.14 ± 3.76 | 60.11 | [53.8-68.56] | 0.91, p=0.34 |
| Lexical Sophistication | 39.51 ± 5.78 | 39.93 | [25-57.94] | 39.01 ± 4.87 | 39.21 | [30.14-47.06] | 0.1, p=0.76 |

NOTE: This table shows both human-coded automated SGT performance between HC and MCI groups. Mean, standard deviation, median, and range are included as well as Kruskal Wallis Chi Square and p-values. Bold text indicates statistically significant differences.

Results of automated scoring also are shown in Table 3. As shown, participants in the MCI group generated more pronouns and a higher pronoun-noun ratio. The groups did not differ in lexical diversity, lexical density, or lexical sophistication.

### Aim 2: Relations between SGT scores and cognitive tests

As shown in Table 4, the episodic memory composite score was significantly correlated with total essential steps, non-essential steps, time to completion, and total words. The direction of these relations showed that participants with better episodic memory abilities generated more words, took more time to complete the script task, and generated more essential and non-essential steps. The strongest relation between episodic memory and SGT essential steps is shown in Figure 1; however, all coefficients suggested small to moderate effect sizes and did not survive FDR correction for multiple comparisons. As for automated measures, the episodic memory composite score was significantly correlated with both pronoun and noun rates, such that participants with better episodic memory had lower pronoun use and higher noun use. A scatterplot of episodic memory and SGT pronoun rate can be seen in Figure 2; however, the significant correlations did not survive FDR correction. The executive function composite score was not significantly associated with any of the human coding or automated measures of the SGT.

**Fig. 1.**
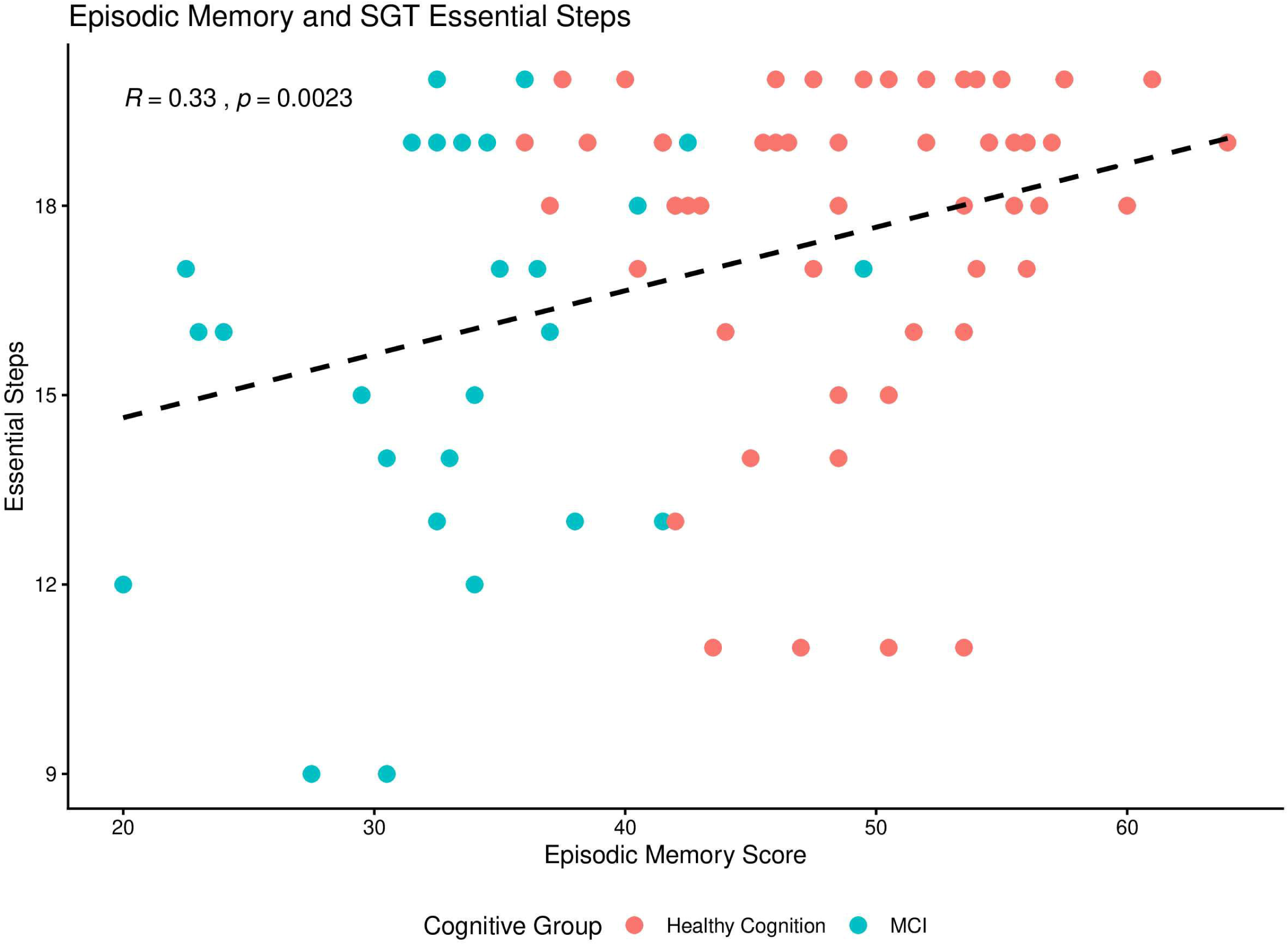

**Fig. 2.**
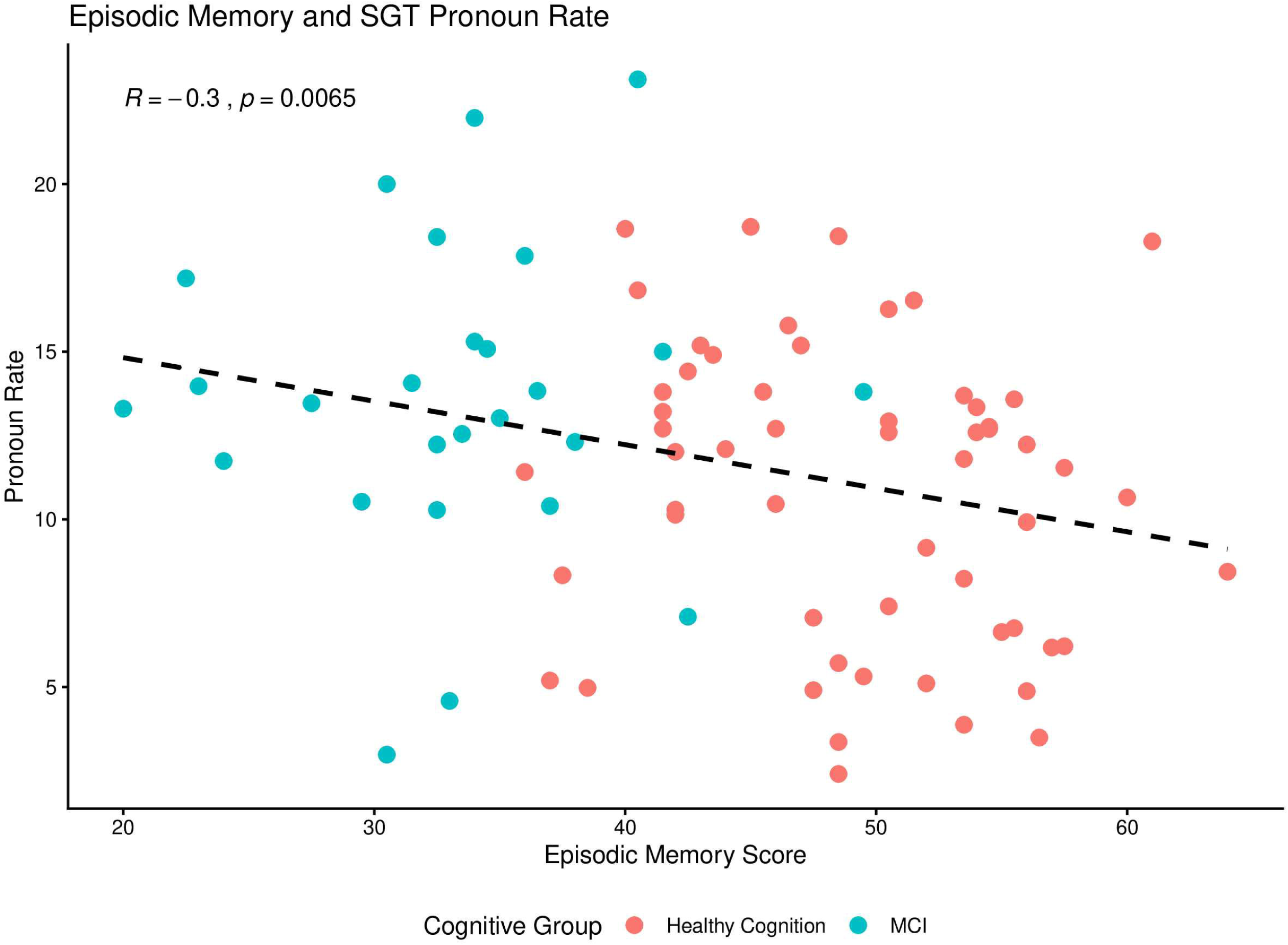

**Table 4:** Aim 2 Spearman Correlation Coefficients; Cognitive Tests and Script Generation.

|  | Episodic Memory Composite<br>Score | Executive Function<br>Composite Score |
| --- | --- | --- |
| Total Essential Steps | **0.33**† | 0.17 |
| Total Non-essential steps | **0.27** * | 0.18 |
| Total Time | **0.29**† | 0.16 |
| Total Words | **0.27** * | 0.21 |
| Total Errors | -0.19 | 0.003 |
| Total Commentary Words | 0.02 | -0.07 |
| Pronoun Rate | **-0.30**† | -0.15 |
| Noun Rate | **0.22** * | 0.1 |
| Pronoun-Noun ratio | **-0.27** * | -0.15 |
| Lexical Diversity | -0.2 | -0.15 |
| Lexical Density | -0.03 | -0.19 |
| Lexical Sophistication | 0.12 | -0.18 |

\*p<0.05; † p<0.01; Bold text indicates significant coefficients, but all significant coefficients did not survive FDR correction.
NOTE: This table reports the correlation coefficients of Spearman Rho values for relations between all human coded and automated script performance scores and episodic memory and executive function composite scores from section 2.3.1.

### Aim 3: Relations between SGT scores and measures of everyday function

Script performance scores were significantly associated with NAT scores (see Table 5). The pattern of correlations showed only SGT human scores were significantly associated with performance on the NAT. Specifically, participants who generated more essential steps on the script task also completed more NAT accomplishment steps, completed the NAT in less time, and made fewer NAT errors. Participants who generated more non-essential steps also took longer to accomplish more NAT steps and made fewer NAT errors. None of the SGT automated scores were significantly associated with the NAT

**Table 5:** Aim 3 Spearman Correlation Coefficients between SGT Scores and Measures of Everyday Function.

|  | NAT steps | NAT time | NAT errors | ECog | FAQ | IADL-C |
| --- | --- | --- | --- | --- | --- | --- |
| Script essential steps | **0.31**<sup>†§</sup> | **-0.26** * | **-0.36**<sup>‡§</sup> | -0.08 | -0.11 | -0.20 |
| Script non-essential steps | **0.37**<sup>†§</sup> | -0.17 | **-0.36**<sup>†§</sup> | -0.14 | -0.13 | **-0.34**<sup>†§</sup> |
| Script time | **0.36**<sup>†§</sup> | 0.03 | **-0.24** * | -0.14 | -0.17 | **-0.28** * |
| Script words | **0.38**<sup>‡§</sup> | -0.15 | **-0.31**<sup>*§</sup> | -0.21 | -0.19 | **-0.34**<sup>†§</sup> |
| Script errors | 0.08 | 0.04 | -0.13 | 0.18 | 0.09 | 0.07 |
| Pronoun Rate | -0.18 | 0.15 | 0.21 | 0.05 | 0.2 | 0.14 |
| Noun Rate | 0.20 | 0.01 | -0.11 | -0.03 | -0.21 | -0.12 |
| Pronoun-Noun Ratio | -0.18 | 0.11 | 0.17 | 0.04 | 0.20 | 0.14 |
| Lexical Diversity | -0.12 | 0.08 | 0.18 | **0.25** * | **0.22** | 0.20 |
| Lexical Density | -0.08 | 0.03 | 0.11 | **0.27** * | 0.14 | **0.23** * |
| Lexical Sophistication | 0.03 | -0.04 | -0.12 | -0.003 | 0.06 | 0.08 |

\*p<0.05; † p<0.01; ‡ p<0.001; § correlation survived FDR correction. Bold text indicates correlations that are significant at p <.05.
NOTE: This table shows Spearman correlation coefficients between SGT measures (human coded and automated scores) and conventional measures of everyday function (NAT performance scores and self-reported questionnaires).

Relations between SGT scores and questionnaires of everyday function showed a different pattern of results. Human scores on the SGT were associated only with the IADL-C questionnaire such that participants who generated more non-essential steps and more words and took more time to complete the SGT also reported greater independence and better daily functioning on the IADL-C. However, automated SGT scores were significantly associated with the IADL-C as well as the FAQ and ECog and the direction of the associations was contrary to expectation. For example, participants who generated scripts with greater lexical diversity and lexical density also reported greater difficulties in everyday function (IADL-C, ECog) and greater functional decline (ECog).

## DISCUSSION

The goal of the current study was to investigate the potential utility of a SGT, that has been previously used to evaluate cognitive function in people with dementia (Roll et al., 2019), to detect mild functional difficulties in people with mild cognitive impairment (MCI). SGT was evaluated using both human coding and automated coding (NLP) to capture a range of performance measures. Our first aim, which was to test differences between participants with MCI versus those with healthy cognition, was supported as participants with MCI generated fewer (essential and non-essential) steps than participants with healthy cognition, even though they did not differ significantly in total words generated. The results of Aim 2 regarding relations between SGT scores and conventional cognitive tests were only partially supported, as SGT measures correlated exclusively with scores on tests of episodic memory and did not significantly correlate with tests of executive function. Finally, analyses supported Aim 3, as performance on the SGT was significantly associated with scores on a performance-based test of everyday function (NAT) and participant’s self-reports of their ability to perform everyday tasks in their daily life (IADLC).

Compared to participants with healthy cognition, participants with MCI produced fewer essential steps on the SGT task and took less time to work on the SGT. These performance differences support the construct validity of the SGT and suggest that people with MCI, who are known to experience mild functional difficulties in everyday life, may have sparce script knowledge or may experience a more rapid decay of script representations. Error rates were relatively low for all participants and did not differ between the groups. Exploratory analyses on automated scores were consistent with the results from human codes, as participants with MCI generated a significantly higher proportion of pronouns and pronoun-noun ratio. A tendency to produce pronouns instead of nouns has been reported as an early marker of AD (Bittner et al., 2024). There is no consensus in the literature regarding whether this shift to pronouns over nouns reflects poor executive function (i.e., perspective taking) or degraded semantic knowledge.

However, our results showed significant associations between SGT measures of noun/pronoun use and scores on tests of episodic memory and not executive function, which may be interpreted as evidence that the noun/pronoun shift is linked to declarative/semantic memory difficulties in people with MCI. Furthermore, “empty speech,” a phenomenon in which cognitively impaired individuals produce nonspecific, vague language has been reported extensively as a marker of neurodegenerative diseases (Ahmed et al., 2013; Eyigoz et al., 2020; Fraser et al., 2015; Mueller et al., 2018; Nicholas et al., 1985; Nyongesa et al., 2025).

Analyses for our second aim regarding cognitive correlates of SGT performance clearly indicated that SGT scores were significantly associated with measures of episodic memory but not executive function. Stronger episodic memory scores were significantly associated with more non-essential steps, longer completion time, and more total words. This finding supports the prediction, based on the Goal-Control model, that knowledge/access of task steps depends on medial temporal lobe processes. However, the prediction, also based on the Goal-Control model (Giovannetti et al., 2021), that SGT errors would be associated with scores on tests of executive function was not supported. This finding could be explained by the relatively few SGT errors, poor sensitivity of the executive function measures included in our study, and/ or the relatively little executive function impairment in our sample; however, the same finding was also observed in a study of people with dementia who did not show such a restricted range in SGT errors and tests of executive function (Roll et al., 2019). Thus, the cognitive processes underlying SGT errors remain poorly understood. Overall, our results indicate that most SGT measures that distinguish participants with MCI versus healthy cognition are strongly associated with episodic memory abilities.

Our third aim was to determine relations between SGT performance and performance of everyday tasks as measured by a validated performance-based test (NAT) (Giovannetti et al., 2019; Seligman et al., 2013). and questionnaires typically used in clinical practice. Results showed that SGT measures of essential and non-essential steps, total words, and completion time were associated with all NAT performance measures, such that participants who generated more script words and steps also accomplished more NAT steps and performed the NAT more quickly and with fewer errors. SGT errors and automated scores were not associated with any of the NAT performance measures. Analyses with self-report questionnaires of everyday function showed SGT measures of non-essential steps and higher generative output (total words) over a longer period (completion time) were associated with higher levels of independence on everyday tasks as measured by the IADL-C.

None of the automated measures were associated with the performance-based test of everyday function (NAT). Automated measures of lexical diversity on the SGT were associated with the ECog and FAQ measures, and lexical density was associated with ECog and IADL-C measures. All associations with automated scores were in the unexpected direction, such that higher lexical diversity and density were associated with lower levels of independence on the respective questionnaire measures. This finding supports prior research on language use and cognitive decline that found that changes in language can be detected before any reports of functional difficulties (Robin et al., 2021). As the questionnaire measures are self-reported, our automated NLP measures of language may have detected changes despite the participant reporting higher independence on the functional activity questionnaires. Further research is needed to interpret this finding.

This study is supported by many strengths, such as its diverse racial demographics. Another strength was that those who transcribed speech samples and conducted human error coding were blinded to conditions-HC or MCI to eliminate bias in human scores. The SGT was brief and easy to administer and scoring of the SGT did not require extensive training in assessment. Additionally, to our knowledge this is the first study to SGT in people with MCI and using automated scoring. Limitations are worth noting. First, the size of the MCI group was relatively small and notably smaller than the healthy cognition group, reducing power and generalizability. Second, older adults who speak ESL were not included in the analytic sample to ensure that SGT measures reflected cognitive abilities rather than linguistic background and culture. Future studies are needed to examine the effects of language acquisition and exposure on SGT measures to increase generalizability to older ESL adults. Future research also should include a more diverse sample of participants regarding years of education and estimated intelligence to improve generalizability.

Our findings suggest that script generation has strong potential as an accessible, inexpensive screening tool for MCI. Script generation provides a highly structured task to measure cognitive abilities that may be especially relevant to everyday functioning. Future research should focus on automated scoring programs that capture essential steps, which may hold the most promise as sensitive SGT measures in large-scale research and in clinical settings as an early detection tool for cognitive and functional decline.

## Data Availability

The dataset analyzed for this study will be available in the Virtual Kitchen Challenge Repository at DOI 10.17605/OSF.IO/MJXWE. In the interim, data requests may be sent to the corresponding author.

## Acknowledgements

This work was supported by the National Institute on Aging (R01AG062503). We would like to acknowledge Emma Pinsky, Adaeze Uwaomah, Daniela McCourt, Giuliana Vallecorsa, Simone Brown, and Sophia Holmqvist for data collection and transcribing audio samples. We also would like to thank Dr. Jamie Reilly for his assistance with coding conceptualization.

## Author Contributions

MR and TG contributed to conceptualization. MR and TG contributed to the methodology. MR, MK, AC, and RC contributed to data collection and implementation. MR and TG contributed to formal analysis. MR and TG contributed to writing (original draft preparation). MK, MM, AC, RC, and TG contributed to writing (review and editing). TG contributed to project administration and funding acquisition. All authors read and agreed to the version of the manuscript intended for publication

