## Supplement C for "Script Generation as an Efficient Measure of Cognition and Everyday Function in Older Adults"

**Script Generation Task Scoresheet**

| **Participant Number:** | |  |  |
| --- | --- | --- | --- |
| **Prepare letter for mailing – essential steps** | |  | **Time** |
|  | write letter |  | Total words |
|  | sign letter |  | Commentary words |
|  | fold letter |  | **Error Analysis** |
|  | put letter in envelope |  | lexical substitution |
|  | stamp on envelope |  | perseveration |
|  | address on envelope |  | sequence error |
|  | return address on envelope |  |  |
|  | seal envelope |  |  |
|  | **TOTAL (8 max)** |  |  |
|  | **Total Non Essential Steps** |  |  |
|  | **Total Steps** |  |  |
| **Make toast- essential steps** | |  | **Time** |
|  | put bread in toaster |  | Total words |
|  | turn toaster on |  | Commentary words |
|  | put butter on toast |  | **Error Analysis** |
|  | put jelly on toast |  | lexical substitution |
|  | **TOTAL (4 max)** |  | perseveration |
|  |  |  | sequence error |
|  | **Total Non Essential Steps** |  |  |
|  | **Total Steps** |  |  |
| **Coffee- essential steps** | |  | **Time** |
|  | heat water |  | Total words |
|  | add grinds/powder |  | Commentary words |
|  | add sugar |  | **Error Analysis** |
|  | add cream |  | lexical substitution |
|  | stir |  | perseveration |
|  | **TOTAL (5 max)** |  | sequence error |
|  | **Total Non Essential Steps** |  |  |
|  | **Total Steps** |  |  |
| **PB & J sandwich – essential steps** | |  | **Time** |
|  | add pb |  | Total words |
|  | add jelly |  | Commentary words |
|  | close sandwich |  | **Error Analysis** |
|  | **TOTAL (3 max)** |  | lexical substitution |
|  |  |  | perseveration |
|  | **Total Non Essential Steps** |  |  |
|  | **Total Steps** |  |  |
| **Lunch box – essential steps** | |  | **Time** |
|  | make sandwich |  | Total words |
|  | wrap sandwich |  | Commentary words |
|  | put sandwich in lunchbox |  | **Error Analysis** |
|  | put snack in lunchbox |  | lexical substitution |
|  | fill thermos |  | perseveration |
|  | seal thermos |  | sequence error |
|  | pack thermos in lunchbox |  |  |
|  | close the lunchbox |  |  |
|  | TOTAL (8 max) |  |  |
|  | **Total Non Essential Steps** |  |  |
|  | **Total Steps** |  |  |
