## Supplement B for "Script Generation as an Efficient Measure of Cognition and Everyday Function in Older Adults"

**Script Generation Scoring Instructions**

1. **Transcribe the recording from the audio file into a word doc.**
   a. Put experimenter’s speech in **bold**, keep participant’s speech unbolded
   b. Put time stamps in parentheses at the beginning and end of participant’s response for each task
   c. Put any notes in *italics*
2. **Go through transcript and use this color key to highlight**:

Commentary - participant comments, examples: “I’m not sure if I said this before”, “Add sugar, that reminds me, I need to stop at the grocery store”

Lexical substitution error - substituting one word for another, example: sticker instead of stamp

Perseveration error - repetition of steps, sub-steps, or words, example: “I would add sugar, then cream, then add sugar”

Sequence error - steps in the wrong order, example: “I’d put the letter in the envelope, then I’d write the letter”

Off task steps - any steps that are unrelated to the task, example: “I’d write the letter, then make myself a cup of tea”

Non-crux steps - any steps related to the task, but not necessary for completion. This includes retrieving objects (ex: get bread from cupboard), opening containers (ex: unwrap butter), and clean-up steps (ex: put peanut butter back)

3. **Enter information into script code sheet**
   a. Use Word’s word count feature (Highlight section you want a word count for, then click on the Review tab > word count)
      i. Get total word count and commentary word count
   b. Record time (end time of participant’s response – start time of participant’s response = total time)
   c. Record number of errors (lexical substitution, perseveration, and sequence errors)
   d. Record steps (task steps (use 1 or 0), off-task, crux) and note step total

Rules/clarifications:

Start time is the beginning of the first thing the participant says after the experimenter finishes giving the prompt, can be a step or commentary/questions.

End time is the end of the last thing the participant says, can be the last step or a declaration that they’re finished

In case of self-correction, include original in error count if applicable (i.e., “Put the toast in the toaster, sorry, the bread in the toaster” would count as one lexical substitution error)

Likewise, if they mention a crux step and then revoke it, they should still get credit for that step (i.e., “put the sugar in the coffee, actually wait, never mind” would still receive credit for ‘add sugar’)

Acknowledgement of a step counts as having completed it (i.e., if participant says, “then I’d put the letter in the addressed envelope”, participant gets accomplishments for put letter in envelope, put address on envelope, put return address on envelope). For the lunchbox task, if the participant says “put the drink in”, they get credit for filling, sealing, and placing the thermos in the lunchbox, since those steps are implied.

Don’t subtract any time for experimenter comments during participant response (experimenters should keep comments brief).

Don’t transcribe filler words (um, uh, like) or add … for pauses (otherwise it will count towards word count).

Don’t count/highlight any errors within commentary.

If the participant mentions an object explicitly (“I’d get out the cream”) and then calls it something else (“I’d pour the milk I just got out into the coffee”), they still get credit for adding cream, but “milk” would count as a lexical substitution error. If they did not mention cream, but just say they’d pour the milk into the coffee, they would not get credit for adding cream, but it would also not be a lexical substitution error and would instead count as a non-crux step.

If the participant includes a juice box or something similar that doesn’t need filling or sealing for the “sandwich, snack, & drink” prompt, they should still get 1s for those task steps, since no omission was made.

If the participant says something like “I’d wrap the sandwich, then I’d make the sandwich, and then wrap the sandwich.” that would count as one sequencing error and one perseveration error, but if the repetition is a correction like “I’d wrap the sandwich, oops, actually I’d make it first and then wrap it”, then they would only get one sequencing error. No perseveration error would be counted since they were repeating the step to correct the sequencing error.

For the “make sandwich” crux step of the lunchbox task, since they can make whatever kind of sandwich they want, do not count the sandwich assembly steps (like toasting the bread, slicing ingredients, placing ingredients on the bread, etc.) as anything, but everything else (retrieving ingredients, opening jars, putting things in the sink) should count as non-crux steps.

If a participant makes a lexical error while describing a crux step, they should still get the accomplishment point for that step, as long as it’s clear what step they’re describing.

RC 4/26: finding someone to eat the sandwich should be counted as non-crux as it is related to the task but they are doing an extra step
