## Supplement A for "Script Generation as an Efficient Measure of Cognition and Everyday Function in Older Adults"

**Everyday Action Script Generation Task Examiner Instructions**

**“I’m going to ask you to tell me all of the steps that you would do to perform an everyday activity. Here’s an example, if I ask you to tell me all of the steps that you would do to brush your teeth, you could say:**

**Open toothpaste**

**Put paste on brush**

**Turn on water**

**Wet toothbrush**

**Put toothbrush in mouth**

**Brush teeth**

**Rinse mouth**

**Rinse teeth**

**Turn off water**

**Do you have any questions?”**

Once the participant understands the task: **“Okay, here’s the first one. Tell me all of the steps that you would do to prepare a letter for mailing.”**

**Stop participant after 3 minutes for each item, and record end time.**

**2. “Tell me all of the steps that you would do to make toast with butter and jelly.”**

**3. “Tell me all of the steps that you would do to prepare instant coffee with cream and sugar.”**

**4. “Tell me all of the steps that you would do to make a sandwich with peanut butter and jelly.”**

**5. “Tell me all of the steps that you would do to prepare a lunchbox with a sandwich, a snack, and a drink.”**

***If the participant has questions, say “Do what you think is best.” Do NOT repeat the instructions.***
